# Closing the Pediatric Divide: A Performance Analysis of the GPT-5 Family in Medical Diagnostics

**DOI:** 10.1101/2025.08.28.25334657

**Authors:** Gianluca Mondillo, Fabio Giovanni Abbate, Mariapia Masino, Simone Colosimo, Alessandra Perrotta, Vittoria Frattolillo

## Abstract

**Background:** Large Language Models (LLMs) have demonstrated significant potential in clinical medicine, but a persistent performance gap exists in the pediatric domain due to its unique complexities. This study provides the first comparative evaluation of the new GPT-5 family (Nano, Mini, and full) to assess the impact of model scale on diagnostic accuracy and this specific adult-pediatric disparity.

**Methods:** A benchmarking study was conducted using 2,000 multiple-choice questions from the MedQA dataset, equally divided between adult (n=1,000) and pediatric (n=1,000) domains. GPT-5, GPT-5 Mini, and GPT-5 Nano were tested via API with standardized parameters (temperature=0, reasoning effort=minimal, verbosity=low, maxtoken=170). Accuracy was calculated and statistically compared across domains for each model.

**Results:** A clear dose-response relationship was observed between model size and accuracy. GPT-5 Nano exhibited a significant performance gap, with an accuracy of 71.0% in adult medicine versus 55.4% in pediatrics (a 15.6 percentage point difference, p<0.001). GPT-5 Mini substantially narrowed this gap to 5.7 points (81.5% vs. 75.8%, p=0.001). Critically, the full GPT-5 model eliminated the disparity, achieving comparable accuracy in adult medicine (86.3%) and slightly higher accuracy in pediatrics (88.5%) (p=0.138). Performance gains from scaling up were disproportionately larger for the pediatric domain.

**Conclusion:** The GPT-5 family marks a substantial advancement in medical AI. The full-size model not only achieves high diagnostic accuracy but, crucially, overcomes the previously documented performance limitations in pediatrics. This demonstrates that sufficient model scale is vital for mastering the nuances of specialized clinical domains. These findings support a tiered implementation strategy based on task criticality and underscore the need for continued validation in real-world clinical settings.

## 1 Introduction

Large Language Models (LLMs) represent one of the most significant innovations in contemporary Artificial Intelligence (AI), based on transformer architectures and trained on vast text corpora that include medical literature, clinical guidelines, and case studies [1]. These models have demonstrated emergent capabilities in clinical reasoning, interpretation of complex symptoms, and diagnostic decision support, opening new perspectives for the application of AI in the healthcare setting [2, 3]. The performance of LLMs in medicine has shown promising results in recent years. Recent studies have documented that advanced models like GPT-4 can achieve diagnostic accuracies comparable to or higher than those of specialist physicians in training on standardized exams and structured clinical questions [4, 5]. However, most of these evaluations have focused on general or adult medicine, with relatively few studies specifically dedicated to the pediatric domain. Pediatrics presents unique peculiarities that could significantly influence the diagnostic performance of LLMs. The pathophysiological specificities of the pediatric patient, weight-based pharmacological and laboratory dosages, clinical presentations that are often atypical compared to adults, and the need to consider neurocognitive developmental stages represent elements of additional complexity [6]. A recent study by Mondillo et al. highlighted how several contemporary LLMs, including advanced models like ChatGPT-3.5, Gemini 2.0, and Claude 3.5 Sonnet, show significantly reduced performance on pediatric questions compared to those in adult medicine, with accuracy differences of up to 10 percentage points for some models [7]. This systematic gap between adult and pediatric medicine suggests intrinsic limitations in the understanding of pediatric specificities by current LLMs. The GPT-5 family from OpenAI [8] represents the most recent evolution of this technology, introducing substantial improvements in reasoning capabilities and architectures optimized for specialized tasks. The availability of three size variants (GPT-5, Mini, Nano) offers a unique opportunity to investigate the relationship between computational complexity and diagnostic accuracy, particularly in the comparison between adult and pediatric medicine. The objective of this study is to comparatively evaluate the diagnostic accuracy of the three GPT-5 variants on a large dataset of clinical questions, with a particular focus on identifying any performance differences between adult and pediatric medicine and the patterns of improvement correlated with model size.

## 2 Materials and Methods

A benchmarking study was conducted to evaluate the diagnostic accuracy of GPT-5, GPT-5 Mini, and GPT-5 Nano on structured clinical questions from the MedQA dataset [9, 10]. From the full dataset, a total of 2000 multiple-choice questions with a single correct answer were randomly extracted, equally distributed between adult medicine (n=1000) and pediatrics (n=1000), and stratified by medical specialties: Allergology, Cardiology, Dermatology, Emergency Medicine, Endocrinology, Gastroenterology, Genetics, Gynecology, Hematology, Immunology, Infectious Diseases, Nephrourology, Neurology, Oncology, Ophthalmology, Orthopedics, Otolaryngology, Pneumology, Psychiatry (Child Psychiatry for pediatrics), Rheumatology, and Surgery. All models were tested via API, sending one question at a time, using standardized parameters: temperature=0 to ensure deterministic outputs, reasoning effort=minimal to reduce response latency by testing the models’ minimum capabilities, verbosity=low to obtain concise and focused answers and max tokens=170 to constrain output length. Accuracy was calculated as the percentage of correct answers out of the total for each model and domain. The 95% confidence intervals for total accuracy were calculated using the Clopper–Pearson method, while the Wilson method was applied for the per-specialty analyses. Differences between adult and pediatric medicine were assessed using the chi-square test (*α <* 0.05). Effect size was quantified with Cramér’s V, and odds of providing a correct response in pediatrics relative to adults were estimated with 95% confidence intervals. Accuracy deltas between the two domains were calculated with their respective confidence intervals using the Newcombe–Wilson method. Confusion matrices, alluvial plots, and radar charts were generated for result visualization.

## 3 Results

Table 1 summarizes the overall accuracy and statistical parameters for all three GPT-5 models. GPT-5 Nano showed an accuracy of 71.0% (95%CI: 68.1%-73.8%) in adult medicine and 55.4% (95%CI: 52.3%-58.5%) in pediatrics, with a difference of 15.6 percentage points (95%CI: -19.8%; -11.4%, p<0.001, Cramér’s V=0.16, OR=0.50, 95%CI: 0.42-0.61). GPT-5 Mini demonstrated an accuracy of 81.5% (95%CI: 79.0%-83.9%) in adult medicine and 75.8% (95%CI: 73.0%-78.4%) in pediatrics, with a difference of 5.7 percentage points (95%CI: -9.3%; -2.1%, p=0.001, Cramér’s V=0.07, OR=0.71, 95%CI: 0.57-0.88). GPT-5 achieved an accuracy of 86.3% (95%CI: 84.0%-88.4%) in adult medicine and 88.5% (95%CI: 86.4%-90.4%) in pediatrics, with a difference of +2.2 percentage points in favor of pediatrics (95%CI: -0.7%; +5.1%, p=0.138, Cramér’s V=0.03, OR=1.22, 95% CI: 0.937-1.59).

**Table 1:** Comparison of GPT-5 models’ accuracy across adult and pediatric domains, with odds ratios (OR) and 95% confidence intervals.

| Model | Accuracy (%) | | $\Delta$ Difference | p-value | Cramér's V | OR (95% CI) |
| --- | --- | --- | --- | --- | --- | --- |
|  | Adult | Pediatric |  |  |  |  |
| GPT-5 Nano | 71.0<br>(68.1–73.8) | 55.4<br>(52.3–58.5) | -15.6 (-19.8;<br>-11.4) | <0.001 | 0.16 | 0.50<br>(0.42–0.61) |
| GPT-5 Mini | 81.5<br>(79.0–83.9) | 75.8<br>(73.0–78.4) | -5.7 (-9.2;<br>-2.1) | 0.001 | 0.07 | 0.71<br>(0.57–0.88) |
| GPT-5 | 86.3<br>(84.0–88.4) | 88.5<br>(86.4–90.4) | +2.2 (-0.7;<br>+5.1) | 0.138 | 0.03 | 1.22<br>(0.94–1.59) |

In adult medicine, GPT-5 showed accuracy above 80% in 18 of the 21 specialties. The highest performances were observed in Allergology (100%), Emergency Medicine (100%), and Otolaryngology (100%). The most challenging specialties were Ophthalmology (58.3%) and Orthopedics (71.4%). In pediatrics, GPT-5 maintained accuracies above 90% in 6 specialties: Allergology (100%), Child Psychiatry (95.6%), Surgery (100%), Pneumology (95.1%), Orthopedics (94.4%), and Otolaryngology (90.5%). In pediatrics, GPT-5 Nano showed deficient performance in Immunology (41.5%), Nephrourology (45.9%), and Hematology (46.1%). See Supplementary Appendix for detailed tables on metrics per specialty.

The alluvial plots (Figures 1 and 2) showed that in the transition from GPT-5 Nano to GPT-5 Mini, an improvement of 10.5% was observed for adult medicine and 20.4% for pediatrics. In the transition from GPT-5 Mini to GPT-5, the improvement was 4.8% for adult and 12.7% for pediatric. The radar charts (Figures 3 and 4) show that GPT-5 maintains consistent performance across all specialties, while GPT-5 Nano exhibits marked variability, which is particularly pronounced in pediatrics. Figures 5, 6, and 7 show the confusion matrices for each model.

**Figure 1.**
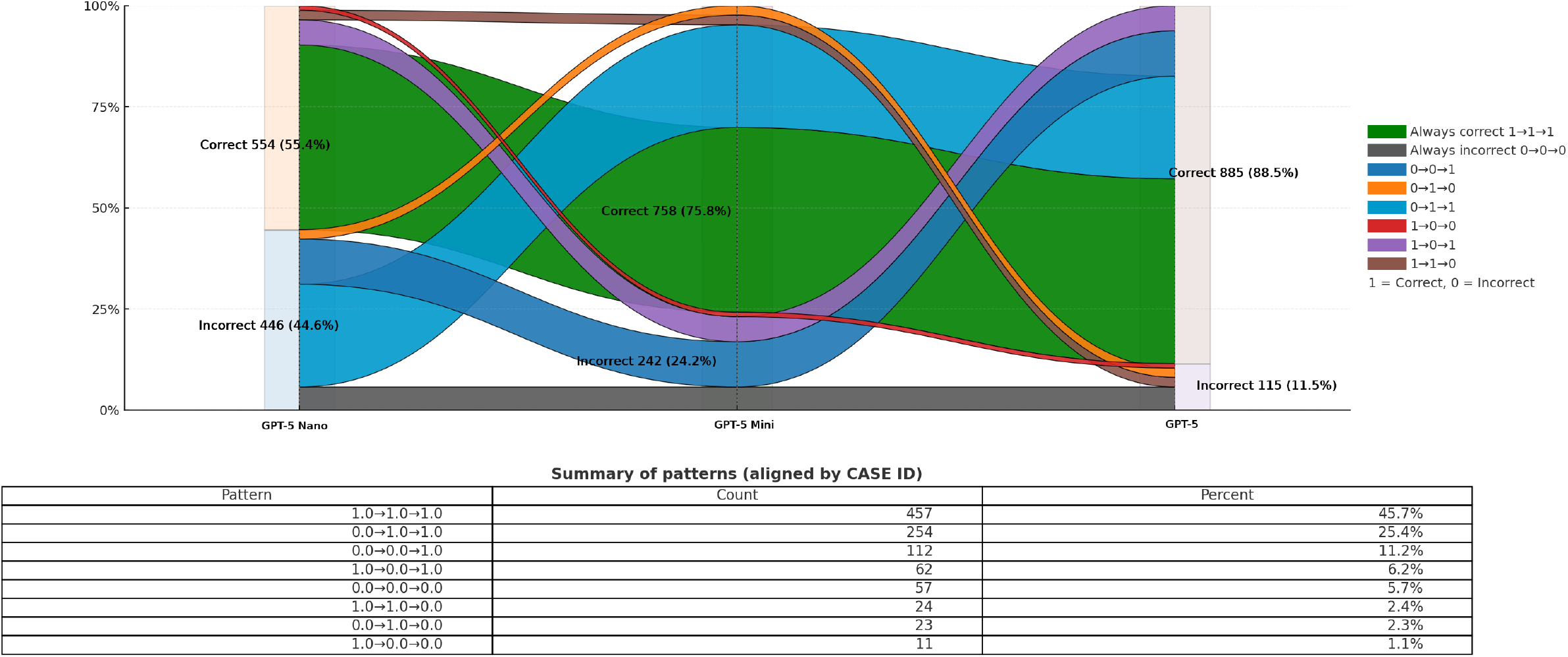
Alluvial Plot for Pediatric questions, displayed on a landscape page for enhanced readability.

**Figure 2.**
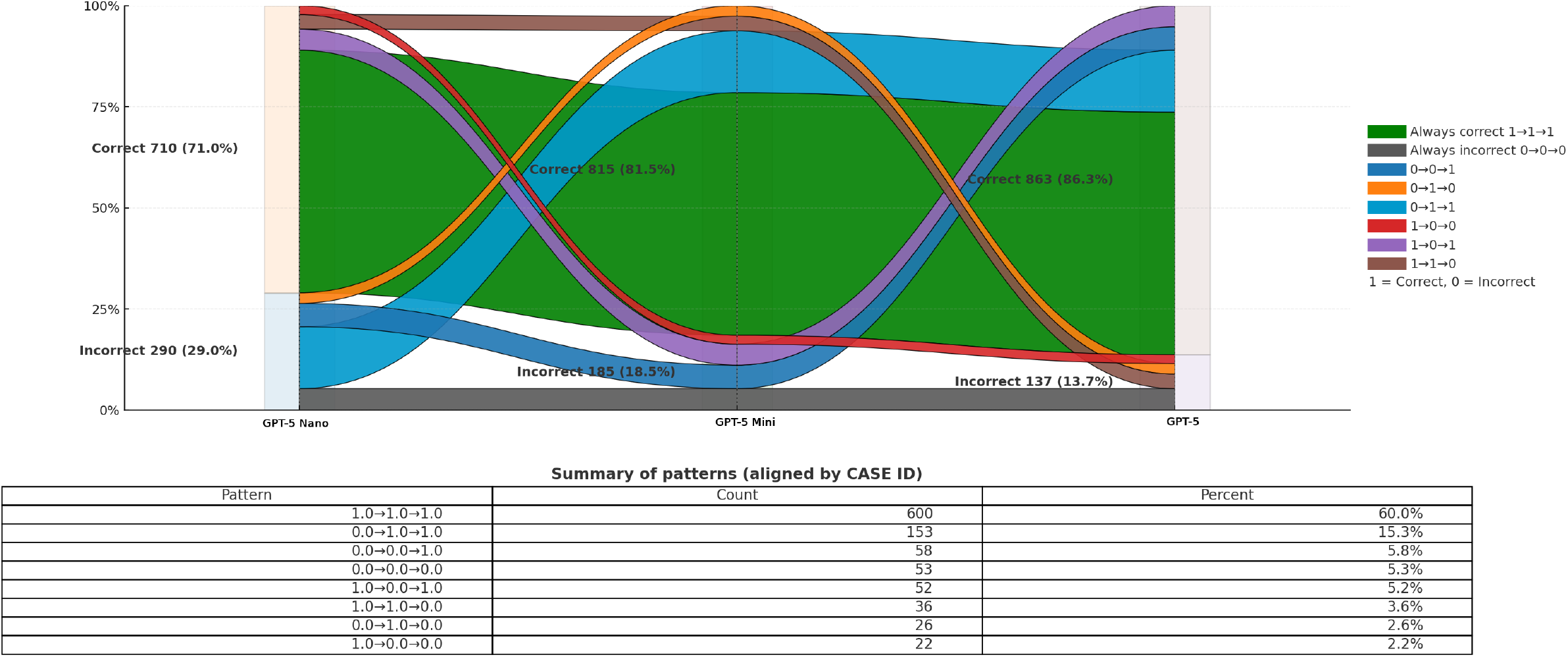
Alluvial Plot for Adult medicine questions, displayed on a landscape page for enhanced readability.

**Figure 3.**
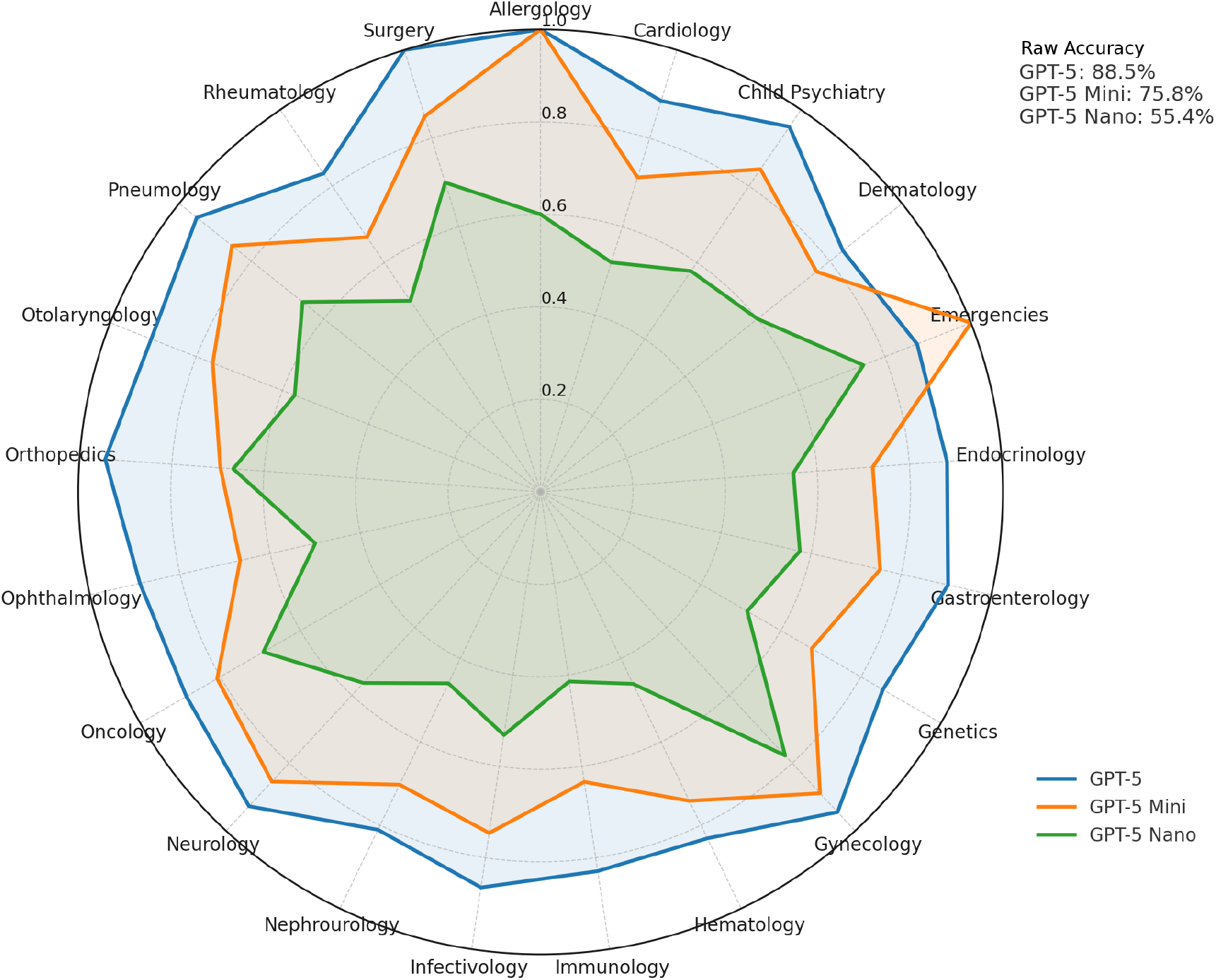
Radar chart for pediatric questions comparing GPT-5 Nano, GPT-5 Mini, and GPT-5.

**Figure 4.**
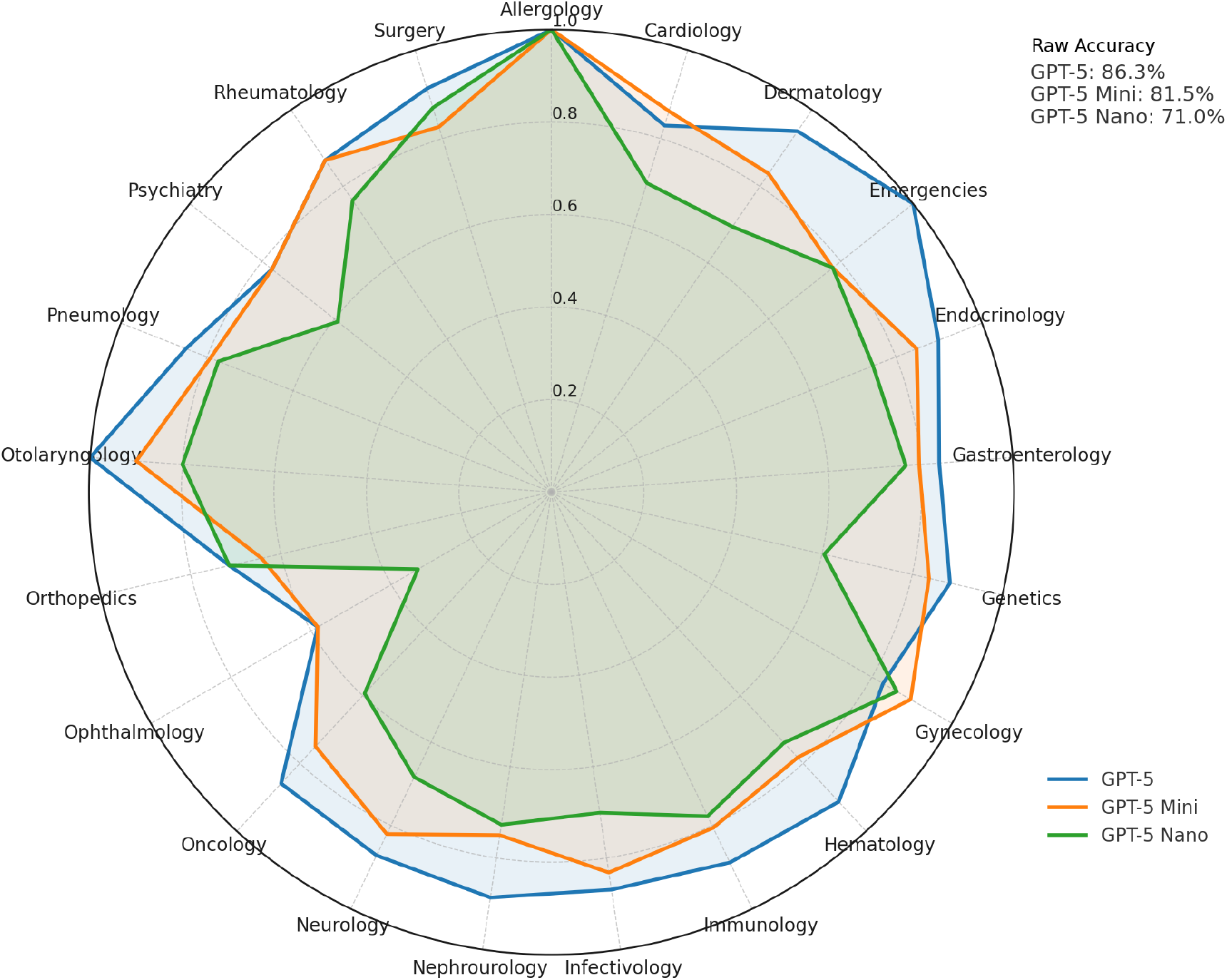
Radar chart for adult questions comparing GPT-5 Nano, GPT-5 Mini, and GPT-5.

**Figure 5.**
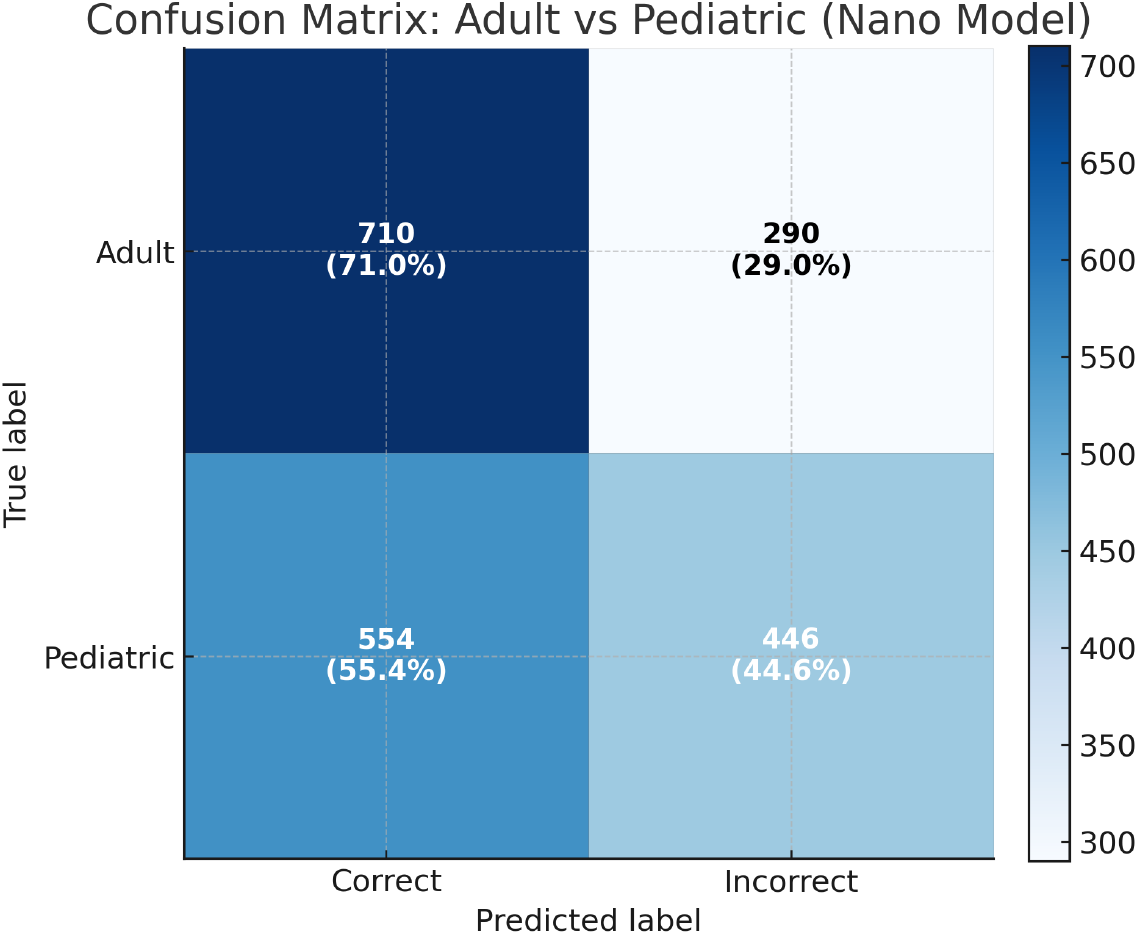
Confusion matrix pediatric vs adult on GPT-5 Nano responses.

**Figure 6.**
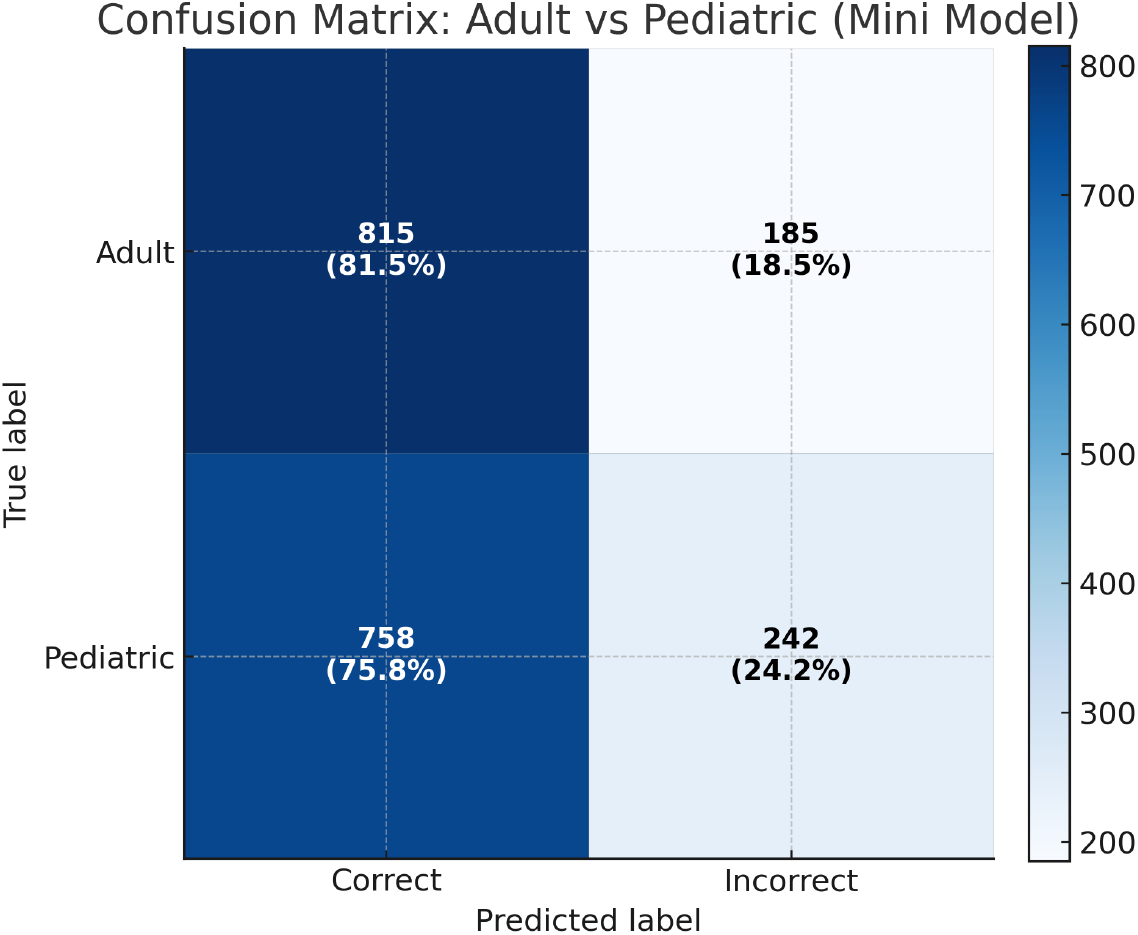
Confusion matrix pediatric vs adult on GPT-5 Mini responses.

**Figure 7.**
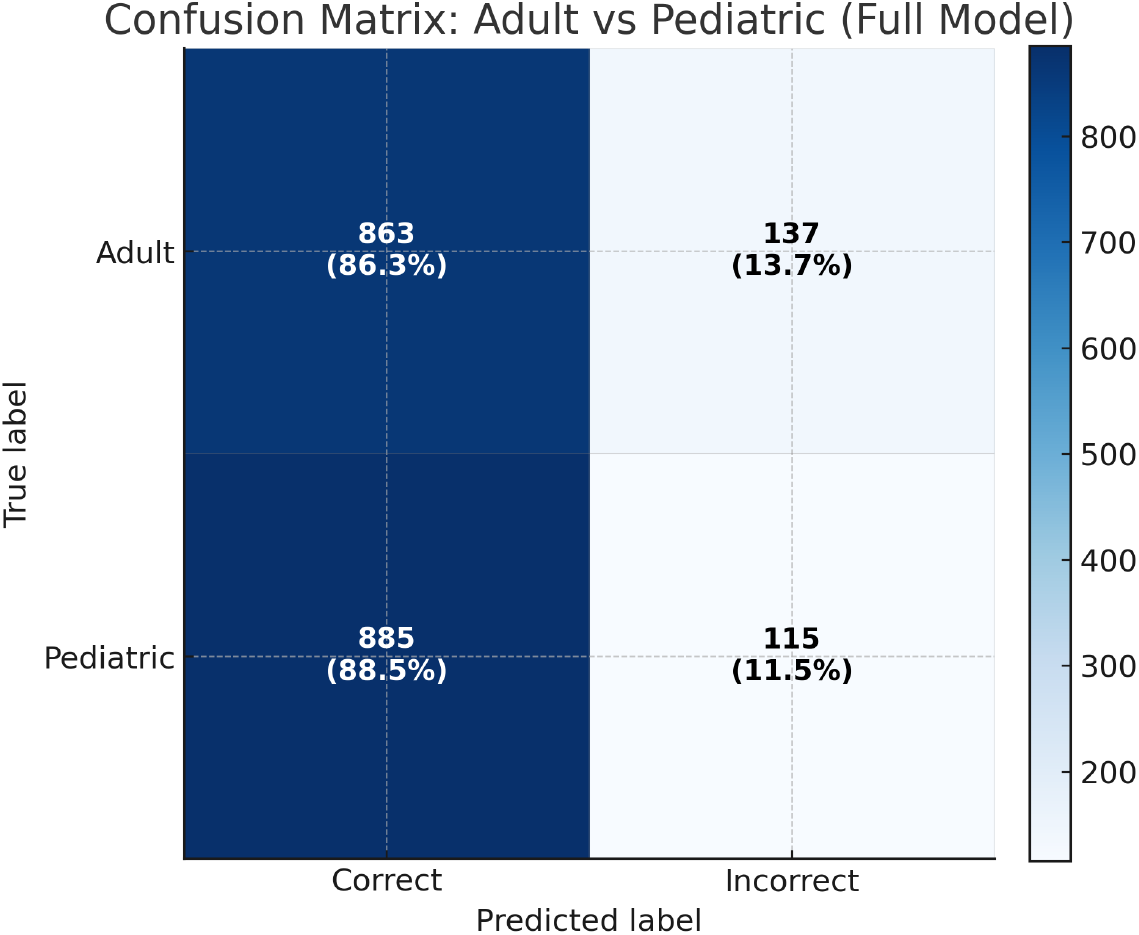
Confusion matrix pediatric vs adult on GPT-5 responses.

## 4 Discussion

The results of this study reveal substantial progress in the performance of GPT-5 models in the medical field, with significant implications for the clinical implementation of diagnostic artificial intelligence systems. The observed dose-response relationship between model size and diagnostic accuracy confirms the importance of computational complexity in clinical reasoning, showing a progressive improvement from GPT-5 Nano (55.4-71.0%) to GPT-5 Mini (75.8-81.5%) to GPT-5 (86.3-88.5%), a pattern that is particularly significant when compared with previous studies on other LLMs. Nori et al. documented that GPT-3.5 achieves an average accuracy of 53.61%, while GPT-4 reaches an average accuracy of 86.7% on the United States Medical Licensing Examination (USMLE), surpassing the exam’s passing threshold by over 20 points [11], a result comparable to the performance of GPT-5 in our study. A more recent study by Bicknell et al. on GPT-4o showed an accuracy of 90.4% vs 81.1% for GPT-4 vs 60.0% for GPT-3.5 on 750 USMLE questions, with medical students achieving an average accuracy of 59.3% [12]. In the study by Kung et al. [13], the version of ChatGPT based on GPT-3.5 (January 2023) achieved an accuracy between 36.1% and 61.3% on USMLE Steps, close to the passing threshold (60%). A second study by Garabet et al. [14] evaluated GPT-4 on USMLE Step 1 style questions and found an accuracy of 86%. In specific skills, GPT-4 had shown 90% accuracy on medical soft skills vs 62.5% for ChatGPT [15]. The performance of GPT-5 (86.3-88.5%) is positioned between that of GPT-4 (86.7%) and GPT-4o (90.4%), suggesting that the incremental improvement in the GPT family continues, but with smaller magnitudes than previous generational leaps. Particularly significant is the comparison with specialized diagnostic capabilities, where GPT-4o showed a diagnostic accuracy of 92.7% and management accuracy of 88.8% [12], values higher than the average performance of GPT-5 but comparable to its performance in the best specialties. The second critical aspect concerns the progressive reduction of the gap between adult and pediatric medicine correlated with model size. GPT-5 Nano shows a marked disparity with 71.0% accuracy for adults versus 55.4% for pediatrics (a difference of 15.6 points, p<0.001, Cramér’s V=0.16). GPT-5 Mini significantly reduces this gap to 5.7 points (81.5% vs 75.8%, p=0.001, Cramér’s V=0.07), while GPT-5 completely eliminates the disparity (+2.2 points in favor of pediatrics, p=0.138). This pattern represents a substantial advancement compared to previous evaluations, in which several less recent generation LLMs showed significantly lower performance on pediatric questions compared to adult ones. In the same study, the most recent models instead showed a substantial reduction of this gap: ChatGPT-4o achieved almost perfect parity between the two domains (83.57% for adult and 83.18% for pediatric, p=0.80) [7]. This result is now surpassed by GPT-5, which shows a slight, though not statistically significant, superiority in pediatrics. This finding is consistent with independent pediatric-focused evaluations, such as an analysis on 500 pediatric MedQA questions that reported an accuracy of 92.8% for ChatGPT O1 and 87.0% for DeepSeek-R1 [16].

The alluvial plots reveal interesting patterns in the transitions between models. The transition from GPT-5 Nano to GPT-5 Mini shows a greater improvement in pediatrics (20.4%) compared to adult medicine (10.5%), as does the subsequent transition from GPT-5 Mini to GPT-5 (12.7% pediatric vs 4.8% adult). This suggests that the increase in model size yields disproportionately greater benefits for understanding pediatric specificities. The radar charts highlight distinctive patterns by specialty. In pediatrics, GPT-5 maintains excellent and consistent performance across all specialties, whereas GPT-5 Nano shows marked irregularities, particularly in Immunology (41.5%), Nephrourology (45.9%), and Hematology (46.1%). Some specialties represent consistent challenges for all models. Ophthalmology shows an accuracy of 58.3% even for GPT-5 in adults, likely due to its highly visual and specialized nature. The choice of API parameters merits discussion. The parameter temperature=0 ensures deterministic outputs, appropriate for reproducibility, though higher temperatures can improve creativity [17]. The parameter reasoning effort=minimal reduces latency by testing minimum capabilities but may have limited the maximum potential, particularly for GPT-5 [18, 19, 20]. The verbosity=low parameter produces concise outputs but may not fully reflect clinical utility where detailed explanations are crucial. For clinical implementation, the results suggest differentiated strategies. GPT-5 represents the optimal choice for critical applications, particularly in pediatrics. GPT-5 Mini offers an acceptable compromise for preliminary screening, and GPT-5 Nano could find application in high-speed scenarios with intensive human supervision. The study’s limitations include the use of multiple-choice questions, which may not reflect real clinical complexity and could be part of the training data. Further limitations are the absence of validation on direct clinical cases and the use of speed-optimized API parameters. Future studies should evaluate performance with higher reasoning effort and on prospective clinical datasets.

## 5 Conclusions

This study provides the first systematic evaluation of the comparative performance of GPT-5 models in adult and pediatric medicine, revealing significant progress in medical artificial intelligence. Performance improves substantially with increasing model size, with GPT-5 achieving accuracy above 85% in both clinical domains. Particularly significant is the elimination of the gap between adult and pediatric medicine with the full model, overcoming limitations documented in previous studies on other LLMs. The analysis of transitions between models demonstrates that the benefits of increased size are greater in pediatrics. Variations across specialties highlight the importance of domain-specific validations before clinical implementation. Practical implementation should consider the trade-off between accuracy and computational resources, with GPT-5 recommended for critical applications and GPT-5 Mini for preliminary screening. Future research should focus on validation in real clinical contexts and on optimization for specific specialties that show sub-optimal performance.

## Data Availability

All data produced in the present study are available upon reasonable request to the authors.

## Supplementary Appendix

### 1. Accuracy for GPT-5

**Table 2:** Accuracy by Specialty (Adult) for GPT-5. Note: the 95% confidence intervals in the table are calculated using the Wilson method.

| Specialty | N | Correct | Accuracy (%) | 95% CI |
| --- | --- | --- | --- | --- |
| Allergology | 4 | 4 | 100.0 | 51.0 – 100.0 |
| Cardiology | 123 | 102 | 82.9 | 75.3 – 88.6 |
| Dermatology | 36 | 34 | 94.4 | 81.9 – 98.5 |
| Emergencies | 9 | 9 | 100.0 | 70.1 – 100.0 |
| Endocrinology | 79 | 71 | 89.9 | 81.3 – 94.8 |
| Gastroenterology | 69 | 58 | 84.1 | 73.7 – 90.9 |
| Genetics | 43 | 38 | 88.4 | 75.5 – 94.9 |
| Gynecology | 29 | 24 | 82.8 | 65.5 – 92.4 |
| Hematology | 46 | 42 | 91.3 | 79.7 – 96.6 |
| Immunology | 36 | 32 | 88.9 | 74.7 – 95.6 |
| Infectivology | 107 | 93 | 86.9 | 79.2 – 92.0 |
| Nephrourology | 44 | 39 | 88.6 | 76.0 – 95.0 |
| Neurology | 117 | 102 | 87.2 | 79.9 – 92.1 |
| Oncology | 64 | 55 | 85.9 | 75.4 – 92.4 |
| Ophthalmology | 12 | 7 | 58.3 | 32.0 – 80.7 |
| Orthopedics | 14 | 10 | 71.4 | 45.4 – 88.3 |
| Otolaryngology | 10 | 10 | 100.0 | 72.2 – 100.0 |
| Pneumology | 53 | 45 | 84.9 | 72.9 – 92.1 |
| Psychiatry | 44 | 34 | 77.3 | 63.0 – 87.2 |
| Rheumatology | 38 | 33 | 86.8 | 72.7 – 94.2 |
| Surgery | 23 | 21 | 91.3 | 73.2 – 97.6 |

**Table 3:** Accuracy by Specialty (Pediatric) for GPT-5. Note: the 95% confidence intervals in the table are calculated using the Wilson method.

| Specialty | N | Correct | Accuracy (%) | 95% CI |
| --- | --- | --- | --- | --- |
| Allergology | 5 | 5 | 100.0 | 56.6 – 100.0 |
| Cardiology | 52 | 46 | 88.5 | 77.0 – 94.6 |
| Child Psychiatry | 45 | 43 | 95.6 | 85.2 – 98.8 |
| Dermatology | 55 | 46 | 83.6 | 71.7 – 91.1 |
| Emergencies | 8 | 7 | 87.5 | 52.9 – 97.8 |
| Endocrinology | 93 | 82 | 88.2 | 80.1 – 93.3 |
| Gastroenterology | 73 | 66 | 90.4 | 81.5 – 95.3 |
| Genetics | 62 | 53 | 85.5 | 74.7 – 92.2 |
| Gynecology | 18 | 17 | 94.4 | 74.2 – 99.0 |
| Hematology | 89 | 74 | 83.1 | 74.0 – 89.5 |
| Immunology | 41 | 34 | 82.9 | 68.7 – 91.5 |
| Infectivology | 126 | 109 | 86.5 | 79.5 – 91.4 |
| Nephrourology | 37 | 30 | 81.1 | 65.8 – 90.5 |
| Neurology | 110 | 102 | 92.7 | 86.3 – 96.3 |
| Oncology | 26 | 23 | 88.5 | 71.0 – 96.0 |
| Ophthalmology | 18 | 16 | 88.9 | 67.2 – 96.9 |
| Orthopedics | 36 | 34 | 94.4 | 81.9 – 98.5 |
| Otolaryngology | 21 | 19 | 90.5 | 71.1 – 97.3 |
| Pneumology | 41 | 39 | 95.1 | 83.9 – 98.7 |
| Rheumatology | 24 | 20 | 83.3 | 64.1 – 93.3 |
| Surgery | 20 | 20 | 100.0 | 83.9 – 100.0 |

### 2. Accuracy for GPT-5 Mini

**Table 4:** Accuracy by Specialty (Adult) for GPT-5 Mini. Note: the 95% confidence intervals in the table are calculated using the Wilson method.

| Specialty | N | Correct | Accuracy (%) | 95% CI |
| --- | --- | --- | --- | --- |
| Allergology | 4 | 4 | 100.0 | 51.0 – 100.0 |
| Cardiology | 123 | 106 | 86.2 | 79.0 – 91.2 |
| Dermatology | 36 | 30 | 83.3 | 68.1 – 92.1 |
| Emergencies | 9 | 7 | 77.8 | 45.3 – 93.7 |
| Endocrinology | 79 | 67 | 84.8 | 75.3 – 91.1 |
| Gastroenterology | 69 | 55 | 79.7 | 68.8 – 87.5 |
| Genetics | 43 | 36 | 83.7 | 70.0 – 91.9 |
| Gynecology | 29 | 26 | 89.7 | 73.6 – 96.4 |
| Hematology | 46 | 36 | 78.3 | 64.4 – 87.7 |
| Immunology | 36 | 29 | 80.6 | 65.0 – 90.2 |
| Infectivology | 107 | 89 | 83.2 | 75.0 – 89.1 |
| Nephrourology | 44 | 33 | 75.0 | 60.6 – 85.4 |
| Neurology | 117 | 96 | 82.1 | 74.1 – 88.0 |
| Oncology | 64 | 48 | 75.0 | 63.2 – 84.0 |
| Ophthalmology | 12 | 7 | 58.3 | 32.0 – 80.7 |
| Orthopedics | 14 | 9 | 64.3 | 38.8 – 83.7 |
| Otolaryngology | 10 | 9 | 90.0 | 59.6 – 98.2 |
| Pneumology | 53 | 42 | 79.2 | 66.5 – 88.0 |
| Psychiatry | 44 | 34 | 77.3 | 63.0 – 87.2 |
| Rheumatology | 38 | 33 | 86.8 | 72.7 – 94.2 |
| Surgery | 23 | 19 | 82.6 | 62.9 – 93.0 |

**Table 5:**
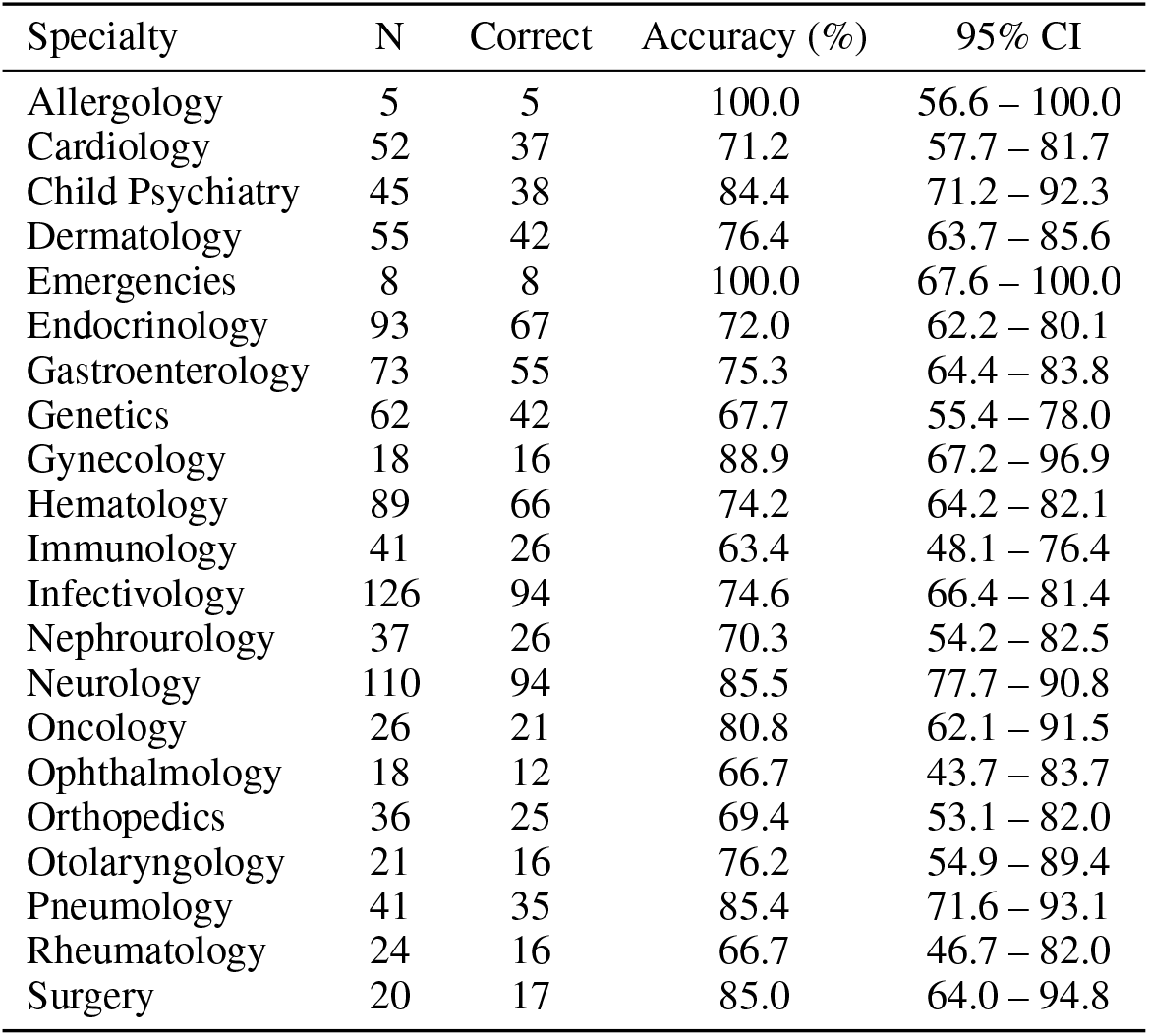
Accuracy by Specialty (Pediatric) for GPT-5 Mini. Note: the 95% confidence intervals in the table are calculated using the Wilson method.

| Specialty | N | Correct | Accuracy (%) | 95% CI |
| --- | --- | --- | --- | --- |
| Allergology | 5 | 5 | 100.0 | 56.6 – 100.0 |
| Cardiology | 52 | 37 | 71.2 | 57.7 – 81.7 |
| Child Psychiatry | 45 | 38 | 84.4 | 71.2 – 92.3 |
| Dermatology | 55 | 42 | 76.4 | 63.7 – 85.6 |
| Emergencies | 8 | 8 | 100.0 | 67.6 – 100.0 |
| Endocrinology | 93 | 67 | 72.0 | 62.2 – 80.1 |
| Gastroenterology | 73 | 55 | 75.3 | 64.4 – 83.8 |
| Genetics | 62 | 42 | 67.7 | 55.4 – 78.0 |
| Gynecology | 18 | 16 | 88.9 | 67.2 – 96.9 |
| Hematology | 89 | 66 | 74.2 | 64.2 – 82.1 |
| Immunology | 41 | 26 | 63.4 | 48.1 – 76.4 |
| Infectivology | 126 | 94 | 74.6 | 66.4 – 81.4 |
| Nephrourology | 37 | 26 | 70.3 | 54.2 – 82.5 |
| Neurology | 110 | 94 | 85.5 | 77.7 – 90.8 |
| Oncology | 26 | 21 | 80.8 | 62.1 – 91.5 |
| Ophthalmology | 18 | 12 | 66.7 | 43.7 – 83.7 |
| Orthopedics | 36 | 25 | 69.4 | 53.1 – 82.0 |
| Otolaryngology | 21 | 16 | 76.2 | 54.9 – 89.4 |
| Pneumology | 41 | 35 | 85.4 | 71.6 – 93.1 |
| Rheumatology | 24 | 16 | 66.7 | 46.7 – 82.0 |
| Surgery | 20 | 17 | 85.0 | 64.0 – 94.8 |

### 3. Accuracy for GPT-5 Nano

**Table 6:** Accuracy by Specialty (Adult) for GPT-5 Nano. Note: the 95% confidence intervals in the table are calculated using the Wilson method.

| Specialty | N | Correct | Accuracy (%) | 95% CI |
| --- | --- | --- | --- | --- |
| Allergology | 4 | 4 | 100.0 | 51.0 – 100.0 |
| Cardiology | 123 | 86 | 69.9 | 61.3 – 77.3 |
| Dermatology | 36 | 25 | 69.4 | 53.1 – 82.0 |
| Emergencies | 9 | 7 | 77.8 | 45.3 – 93.7 |
| Endocrinology | 79 | 59 | 74.7 | 64.1 – 83.0 |
| Gastroenterology | 69 | 53 | 76.8 | 65.6 – 85.2 |
| Genetics | 43 | 26 | 60.5 | 45.6 – 73.6 |
| Gynecology | 29 | 25 | 86.2 | 69.4 – 94.5 |
| Hematology | 46 | 34 | 73.9 | 59.7 – 84.4 |
| Immunology | 36 | 28 | 77.8 | 61.9 – 88.3 |
| Infectivology | 107 | 75 | 70.1 | 60.8 – 77.9 |
| Nephrourology | 44 | 32 | 72.7 | 58.2 – 83.7 |
| Neurology | 117 | 80 | 68.4 | 59.5 – 76.1 |
| Oncology | 64 | 38 | 59.4 | 47.1 – 70.5 |
| Ophthalmology | 12 | 4 | 33.3 | 13.8 – 60.9 |
| Orthopedics | 14 | 10 | 71.4 | 45.4 – 88.3 |
| Otolaryngology | 10 | 8 | 80.0 | 49.0 – 94.3 |
| Pneumology | 53 | 41 | 77.4 | 64.5 – 86.5 |
| Psychiatry | 44 | 26 | 59.1 | 44.4 – 72.3 |
| Rheumatology | 38 | 29 | 76.3 | 60.8 – 87.0 |
| Surgery | 23 | 20 | 87.0 | 67.9 – 95.5 |

**Table 7:** Accuracy by Specialty (Pediatric) for GPT-5 Nano. Note: the 95% confidence intervals in the table are calculated using the Wilson method.

| Specialty | N | Correct | Accuracy (%) | 95% CI |
| --- | --- | --- | --- | --- |
| Allergology | 5 | 3 | 60.0 | 23.1 – 88.2 |
| Cardiology | 52 | 27 | 51.9 | 38.7 – 64.9 |
| Child Psychiatry | 45 | 26 | 57.8 | 43.3 – 71.0 |
| Dermatology | 55 | 33 | 60.0 | 46.8 – 71.9 |
| Emergencies | 8 | 6 | 75.0 | 40.9 – 92.9 |
| Endocrinology | 93 | 51 | 54.8 | 44.7 – 64.6 |
| Gastroenterology | 73 | 42 | 57.5 | 46.1 – 68.2 |
| Genetics | 62 | 32 | 51.6 | 39.4 – 63.6 |
| Gynecology | 18 | 14 | 77.8 | 54.8 – 91.0 |
| Hematology | 89 | 41 | 46.1 | 36.1 – 56.4 |
| Immunology | 41 | 17 | 41.5 | 27.8 – 56.6 |
| Infectivology | 126 | 67 | 53.2 | 44.5 – 61.7 |
| Nephrourology | 37 | 17 | 45.9 | 31.0 – 61.6 |
| Neurology | 110 | 62 | 56.4 | 47.0 – 65.3 |
| Oncology | 26 | 18 | 69.2 | 50.0 – 83.5 |
| Ophthalmology | 18 | 9 | 50.0 | 29.0 – 71.0 |
| Orthopedics | 36 | 24 | 66.7 | 50.3 – 79.8 |
| Otolaryngology | 21 | 12 | 57.1 | 36.5 – 75.5 |
| Pneumology | 41 | 27 | 65.9 | 50.5 – 78.4 |
| Rheumatology | 24 | 12 | 50.0 | 31.4 – 68.6 |
| Surgery | 20 | 14 | 70.0 | 48.1 – 85.5 |

